# Clinicopathologic Predictors of PSMA-PET Detection, Recurrence Pattern, and Biochemical Outcome Following Salvage Radiotherapy After Radical Prostatectomy

**DOI:** 10.64898/2026.10.16.26365643

**Authors:** Ramotallah Jubril, Arpan V. Prabhu, Omar Ishaq, Jesus A. Ocana, Mark Tann, Christopher Njeh, Clinton D. Bahler, Jordan Holmes, Oluwaseyi Oderinde

## Abstract

**Purpose:** Biochemical recurrence (BCR) after radical prostatectomy (RP) presents heterogeneous recurrence patterns and variable outcomes following salvage radiotherapy (sRT). We evaluated clinicopathologic determinants of PSMA-PET/CT detection, anatomical recurrence distribution, and biochemical disease-free survival (bDFS) following PSMA-PET-guided sRT.

**Methods:** This retrospective single-institution study included 140 patients with BCR after RP who underwent ^68^Ga-PSMA-PET/CT prior to sRT with or without androgen deprivation therapy. PSMA-PET-positive findings were categorized as prostate bed only (PBO) or pelvic nodal involvement with or without the prostate bed (PLNI). Receiver operating characteristic (ROC) analysis, logistic regression, and Cox proportional hazards models were employed.

**Results:** PSMA-PET detected locoregional recurrence in 77/140 patients (55%). On multivariable analysis, PSA at recurrence (OR=2.07; p=0.003) and age (OR=1.07; p=0.016) were the sole independent predictors of PSMA-PET positivity. ROC analysis yielded AUC=0.74 (optimal threshold of 0.58 ng/mL). Among PSMA-positive patients, PBO was associated with a longer time to BCR (tBCR) (37.5 versus 15 months; p=0.006). Seminal vesicle invasion (SVI) and pT-stage predicted PLNI on univariable analysis. Among 124 of 140 evaluable patients (median follow-up 31 months), estimated bDFS at 24, 36, and 60 months was 89.7%, 80.2%, and 54.1%, respectively. SVI (HR=2.39; p=0.031), EPE (HR=2.82; p=0.012), pT-stage (HR=1.64; p=0.021), and pN-stage (HR=1.83; p=0.045) were associated with increased failure on univariable Cox regression, though none retained independent significance.

**Conclusion:** PSA burden and age independently predicted PSMA-PET detection. Recurrence patterns reflected disease behavior, as PBO was associated with longer tBCR. Adverse pathologic features were associated with pelvic nodal disease and reduced biochemical control; none remained independently predictive.

## Introduction

Biochemical recurrence (BCR) following radical prostatectomy (RP) represents a critical inflection in prostate cancer (PCa) management occurring in 20-50% of patients within ten years ^1^. Defined by increasing prostate-specific antigen (PSA) level following RP, BCR is indicative of disease progression preceding the development of radiographically evidenced disease with increased risk of metastatic and cancer-specific mortality ^2,3^. Salvage radiotherapy (sRT) delivered to the prostate bed, with or without pelvic nodal irradiation and androgen deprivation therapy (ADT), remains the standard of care for presumed nonmetastatic recurrence and yields durable biochemical control in a meaningful subset of patients ^4–6^. Clinical outcomes after sRT vary widely, reflecting underlying differences in disease burden, spatial distribution of recurrence, and treatment intensification. In recent times, prostate-specific membrane antigen positron emission tomography (PSMA-PET) imaging has been integrated as a sensitive tool enabling detection of recurrent disease at PSA levels where conventional imaging often performs poorly. Prospective and meta-analytic data have consistently demonstrated that ^68^Ga-PSMA-11 PET/CT achieves markedly superior sensitivity and specificity for detection of locoregional (prostate bed and pelvic lymph) and distant recurrence, with detection rate increasing with PSA burden ^7–9^. Beyond diagnosis, PSMA-PET also alters management in a substantial proportion of patients, including men considered for salvage radiotherapy, by changing target volumes, treatment fields, or the decision to intensify therapy ^10–12^.

While PSMA-PET improves disease localization, its integration into salvage decision-making has not fully resolved prognostic uncertainty in men with locoregional recurrence. Prior investigations have largely focused on PSA as the primary driver of PSMA-PET detection, with limited evaluation on how prostatectomy-derived pathological features independently of PSA burden shaped imaging yield and anatomical distribution of locoregional recurrence. More importantly, it remains poorly understood whether clinicopathological risk factors associated with imaging positivity or the PSMA-PET-defined recurrence patterns meaningfully translate into prognostic stratification following PSMA-PET-guided sRT. Accordingly, in a contemporary cohort of men with post-radical BCR staged with ^68^Ga-PSMA-11 PET/CT before salvage radiotherapy, we addressed three pre-specified objectives: (1) to evaluate the clinicopathological determinants of locoregional PSMA-PET detection across the full cohort; (2) to characterize the anatomical distribution of recurrence among PSMA-PET-positive patients; and (3) to assess the prognostic value of clinicopathology and imaging-defined variables for biochemical disease-free survival following PSMA-PET-guided sRT. In doing so, we seek to define the extent to which conventional pathological risk markers retain clinical relevance in the contemporary PSMA imaging era.

## Methods

### Patients Cohort

This retrospective single-center study evaluated patients who underwent RP for confirmed PCa and subsequently developed BCR between 2017 and 2023. BCR was defined as two consecutive prostate-specific antigen (PSA) measurements ≥0.2 ng/mL following undetectable post-operative PSA, in accordance with established consensus criteria ^13^. Eligible patients were required to have undergone RP, met the BCR criteria, undergone ^68^GaPSMA-11-PET-based imaging, and subsequently received sRT with or without concurrent androgen deprivation therapy (ADT). Patients were included irrespective of PSMA-PET findings (positive or negative) to enable comparative analyses of detection, recurrence patterns, and treatment outcomes. Among PSMA-PET-positive patients, anatomical recurrence pattern analyses evaluated only locoregional sites (prostate bed and pelvic lymph nodes). Exclusion criteria were (1) incomplete clinicopathology or biochemical data at BCR; (2) evidence of metastatic disease defined as involvement of recurrence beyond the regional pelvis, including the distant lymph nodes (at or above the common iliac bifurcation), bone, or visceral organs; (3) salvage therapy initiated prior to PSMA-PET imaging; (4) PSA persistence following RP, defined as PSA ≥ 0.1 ng/ml without achieving an undetectable postoperative PSA, and (5) loss of follow-up before documentation of treatment failure. After applying these criteria, 140 patients comprised the final analytic cohort.

Clinical variables extracted included age at BCR, PSA level at recurrence prior to PSMA-PET (BCR-PSA), time to biochemical recurrence (tBCR), and salvage treatment details, including time to secondary biochemical recurrence (tsBCR). Pathologic variables obtained from prostatectomy specimens included International Society of Urological (ISUP) grade (pISUP), T-stage (pT-stage), nodal status (pN-stage), presence of extraprostatic extension (EPE), positive surgical margin status (PSM), and seminal vesicle invasion (SVI).

### PSMA-PET Acquisition and Interpretation

All patients underwent a ^68^Ga-PSMA-PET/CT imaging from vertex to mid-thigh using an integrated whole-body PET/CT system (Biograph Vision 600 PET/CT; Siemens Medical Solutions, Malvern, PA, USA) comprising an early acquisition at 5 minutes post-injection to assess physiological biodistribution followed by a whole-body image acquisition at 60 ± 5 minutes post-administration of ^68^Ga-PSMA-11 to optimize lesion detection. All scans were independently reviewed and interpreted by board-certified nuclear medicine physicians as previously described ^14^. PSMA-PET positive scans were defined by suspicious tracer uptake exceeding background activity that was not attributed to physiological distribution and was consistent with disease recurrence. For PSMA-PET-positive scans, recurrence sites were categorized by anatomical location as: (1) prostate bed only (PBO) or (2) pelvic lymph node involvement with or without prostate bed disease (PLNI). Pelvic lymph nodes were nodes below the common iliac bifurcation. Patients without suspicious uptake were classified as PSMA-PET negative.

### Salvage Radiotherapy and Follow-up

Salvage radiotherapy was delivered according to institutional standards of care, incorporating PSMA-PET findings informing treatment where lesions were identified. All patients received intensity-modulated image-guided therapy with target volume definition, delivery dose, and use of concurrent ADT determined at the discretion of the treating radiation oncologist. Patients were followed longitudinally with serial serum PSA measurements performed at regular intervals per institutional clinical practice. Secondary biochemical failure (sBCR) following sRT was defined using modified Nara criteria as: (1) PSA decline to < 0.2 ng/mL after sRT, followed by a confirmed rise to ≥0.2 ng/mL on two consecutive measurements or (2) PSA level rise without a decline to PSA <0.2 ng/mL after sRT ^15^.

Time-to-event outcomes were calculated from the start of salvage radiotherapy to the date of biochemical failure or last follow-up (censoring). Among the 140 patients included, 16 had incomplete time-to-event data point, restricting the bDFS analysis to 124 patients.

### Statistical analysis

Three pre-specified analyses were evaluated: (1) PSMA detection assessed across all 140 patients; (2) anatomical recurrence localization (PBO versus PLNI) evaluated among PSMA-PET-positive patients; and (3) bDFS following sRT, measured from the first day of salvage radiotherapy to sBCR or last follow-up. Continuous variables were summarized as mean (±SD; standard deviation) or median (IQR; interquartile range) according to distribution assessed with the Shapiro-Wilk test. Categorical variables were reported as frequencies and percentages.

Comparisons between PSMA-PET-negative and positive patients were performed using the t-test or Mann-Whitney U test for continuous data and chi-square test or Fisher’s test for categorical variables as appropriate. Overall diagnostic accuracy of PSMA-PET relative to PSA burden at recurrence was assessed using receiver operating characteristics (ROC) curve analysis with area under the curve (AUC) and 95% confidence interval (95% CI) reported. The optimal PSA threshold was determined by maximizing the Youden index. Among PSMA-PET-positive patients, similar comparative analyses were conducted to evaluate associations between clinicopathologic factors and anatomical recurrence patterns (PBO and PLNI).

Univariable logistic regression (LR) analyses were first performed to identify predictors of: (1) PSMA-PET detection and (2) recurrence distribution pattern among PSMA-positive patients. Variables demonstrating associations at p <0.05 on univariable analysis were subsequently entered into multivariable LR models to identify independent predictors. Odds ratios (OR) with 95% CI were reported.

For sRT outcomes, time-to-event outcomes were analyzed using Kaplan-Meier methodology with log-rank tests for between-group comparisons. Median survival times with 95% confidence intervals were estimated when sufficient events occurred. Cox proportional hazards regression identified predictors of biochemical disease-free survival (bDFS) following sRT, with hazard ratios (HR) and 95% CI reported. All statistical tests were two-sided, with a p<0.05 considered statistically significant. Analysis was performed using Python (version 3.9) and SPSS (IBM Corp., Armonk, NY).

## Results

### Patients Characteristics

A total of 140 patients with BCR following RP who underwent ^68^Ga-PSMA-PET/CT imaging prior to sRT were included in the analysis. The baseline characteristics of the entire cohort are detailed in **Table 1**. The mean age at BCR was 67.1 years (SD: 7.2). The median tBCR from RP was 24 months (IQR: 7.75-56.25), with 88 patients (62.9%) experiencing early recurrence (≤36 months) and 52 patients (37.1%) demonstrating delayed recurrence (>36 months). The median BCR-PSA was 0.57 ng/mL (IQR, 0.35-1.19).

**Table 1:**
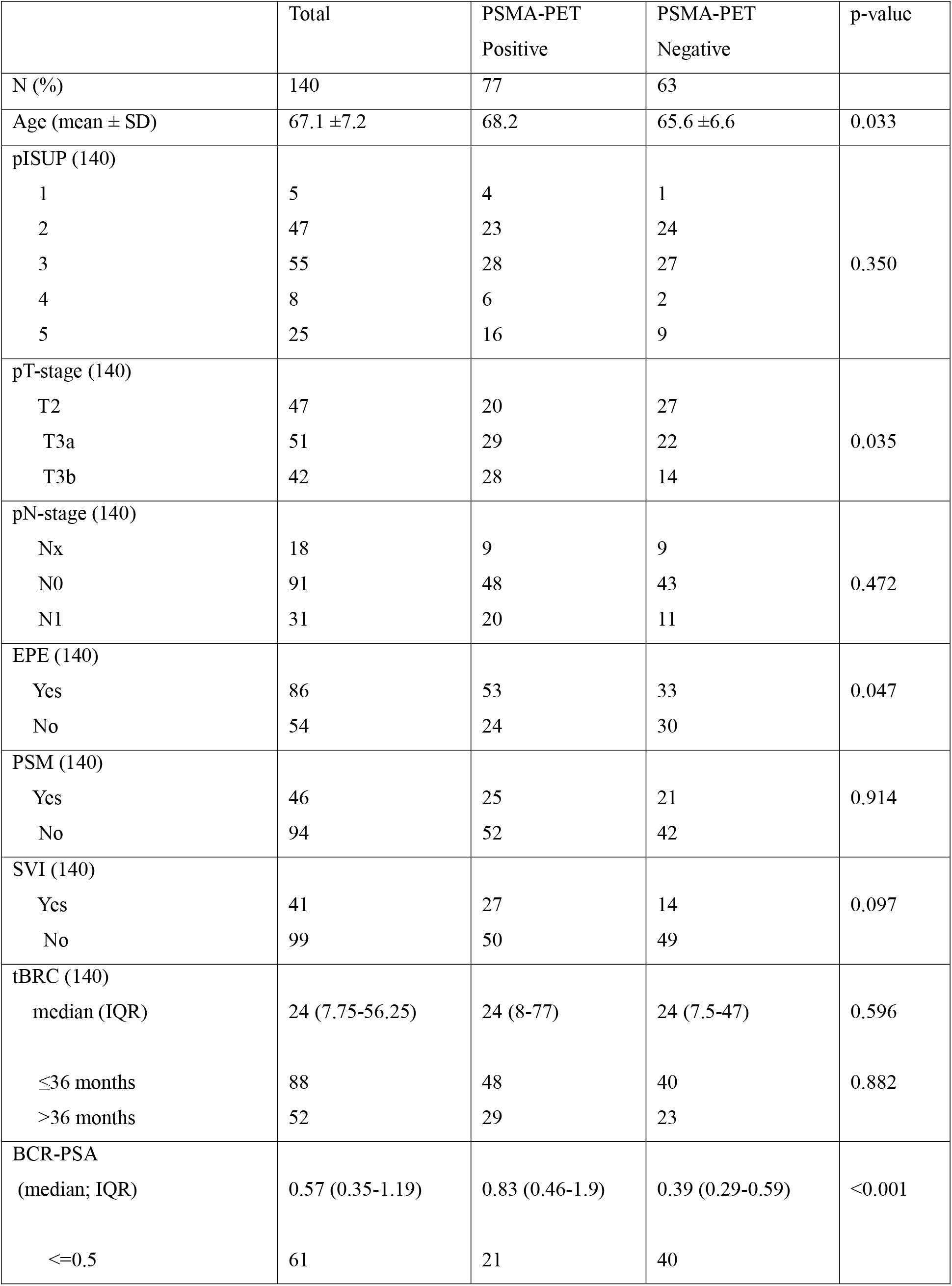
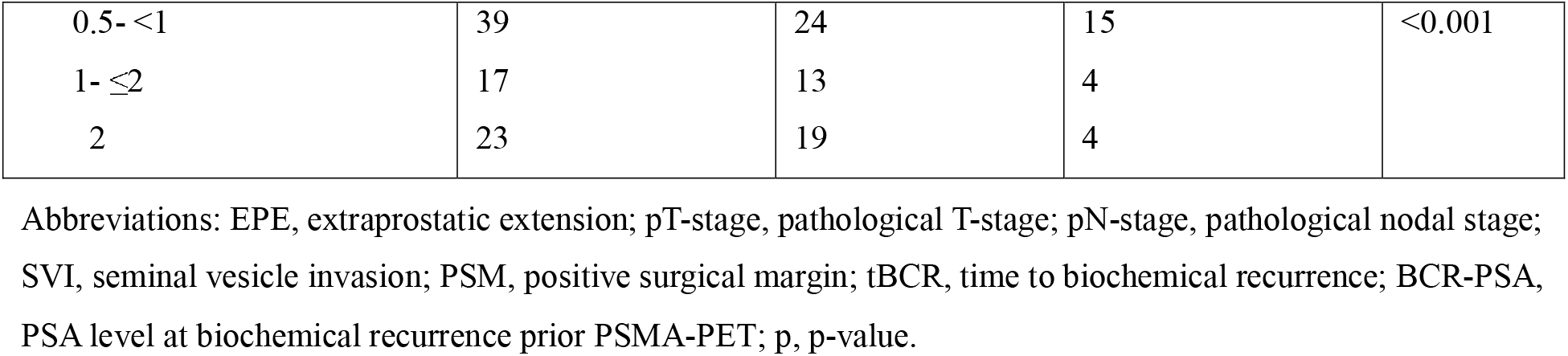
General patient characteristics investigated. Characteristics stratified by PSMA-PET detection.

|  | Total | PSMA-PET<br>Positive | PSMA-PET<br>Negative | p-value |
| --- | --- | --- | --- | --- |
| N (%) | 140 | 77 | 63 |  |
| Age (mean ± SD) | 67.1 ± 7.2 | 68.2 | 65.6 ± 6.6 | 0.033 |
| pISUP (140) |  |  |  |  |
| 1 | 5 | 4 | 1 | 0.350 |
| 2 | 47 | 23 | 24 |  |
| 3 | 55 | 28 | 27 |  |
| 4 | 8 | 6 | 2 |  |
| 5 | 25 | 16 | 9 |  |
| pT-stage (140) |  |  |  |  |
| T2 | 47 | 20 | 27 | 0.035 |
| T3a | 51 | 29 | 22 |  |
| T3b | 42 | 28 | 14 |  |
| pN-stage (140) |  |  |  |  |
| Nx | 18 | 9 | 9 | 0.472 |
| N0 | 91 | 48 | 43 |  |
| N1 | 31 | 20 | 11 |  |
| EPE (140) |  |  |  |  |
| Yes | 86 | 53 | 33 | 0.047 |
| No | 54 | 24 | 30 |  |
| PSM (140) |  |  |  |  |
| Yes | 46 | 25 | 21 | 0.914 |
| No | 94 | 52 | 42 |  |
| SVI (140) |  |  |  |  |
| Yes | 41 | 27 | 14 | 0.097 |
| No | 99 | 50 | 49 |  |
| tBRC (140) |  |  |  |  |
| median (IQR) | 24 (7.75-56.25) | 24 (8-77) | 24 (7.5-47) | 0.596 |
| ≤36 months | 88 | 48 | 40 | 0.882 |
| >36 months | 52 | 29 | 23 |  |
| BCR-PSA<br>(median; IQR) | 0.57 (0.35-1.19) | 0.83 (0.46-1.9) | 0.39 (0.29-0.59) | <0.001 |
| ≤0.5 | 61 | 21 | 40 | |
| 0.5- <1 | 39 | 24 | 15 | <0.001 |
| 1- ≤2 | 17 | 13 | 4 |  |
| 2 | 23 | 19 | 4 |  |

Abbreviations: EPE, extraprostatic extension; pT-stage, pathological T-stage; pN-stage, pathological nodal stage; SVI, seminal vesicle invasion; PSM, positive surgical margin; tBCR, time to biochemical recurrence; BCR-PSA, PSA level at biochemical recurrence prior PSMA-PET; p, p-value.

### Determinant of PSMA-PET Detection

^68^Ga-PSMA-11 PET/CT detected locoregional recurrence in 77 of 140 patients (55%). PSMA-PET-positive patients were older than those with negative scans (mean age of 68.2 versus 65.6 years; p=0.033). Notably, BCR-PSA levels were markedly higher in the PSMA-PET-positive group (median 0.83 ng/mL [IQR, 0.46-1.9] versus 0.39 ng/mL [IQR, 0.29-0.59]; p<0.001) with PSMA-PET positivity increasing progressively across the PSA strata (p <0.001). Pathological features associated with PSMA-PET positivity included higher pT-stage (p=0.035) and EPE (p=0.047), whereas pISUP grade, pN-stage, margin status, and SVI did not differ significantly between detection groups. Comparative characteristics are presented in **Table 1**.

On univariable LR analysis, age at BCR (OR = 1.053; 95% CI = 1.004-1.106; p= 0.034), BCR-PSA (OR= 2.022; 95% CI= 1.234-3.313; p= 0.005), pT-stage (OR = 1.649; 95% CI= 1.070-2.539; p= 0.0233), and EPE (OR= 2.008; 95% CI= 1.006 - 4.005; p= 0.048) were associated with PSMA-PET positivity. However, in multivariate analysis, only BCR-PSA (OR= 2.07; 95% CI= 1.27-3.39; p=0.003) and age at BCR (OR= 1.07; 95% CI= 1.01-1.13; p=0.016) remained independently significant.

ROC analysis for BCR-PSA level demonstrated an AUC of 0.74 (95% CI, 0.65-0.81; p<0.001) (**Fig. 1**). The optimal BCR-PSA threshold maximizing the Youden index was 0.58 ng/ml, achieving a sensitivity of 68% and a specificity of 75% for predicting PSMA-PET positivity.

**Fig 1:**
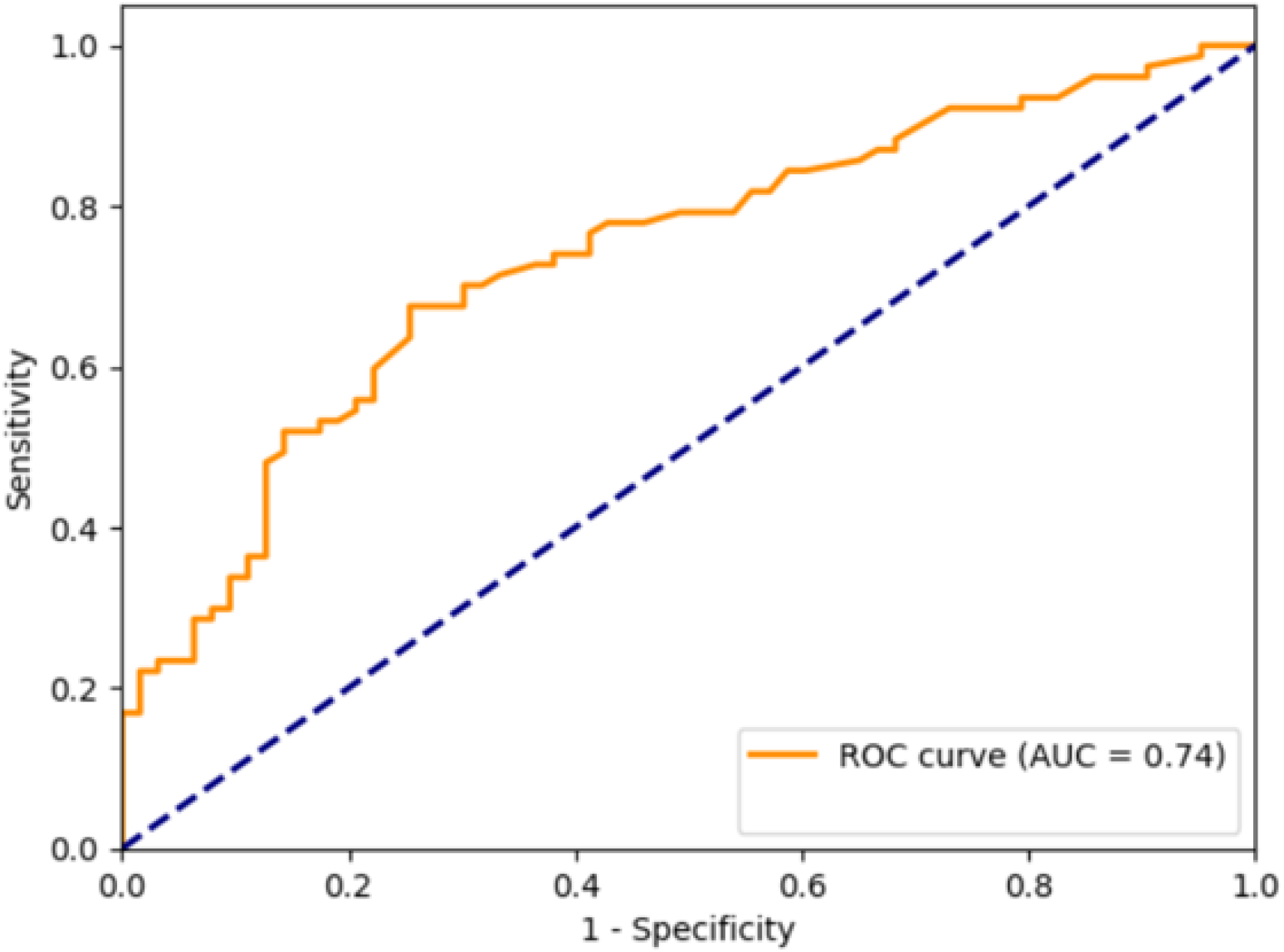
Receiver operating characteristic curve for BCR-PSA predicting PSMA-PET detection. Abbreviations: AUC, area under the curve; ROC, receiver operating characteristic.

### Anatomical Recurrence Pattern Among PSMA-Positive Patients

Among the 77 patients with PSMA-PET-positive, 28 (36.4%) had PBO and 49 patients (63.6%) had PLNI (**Table 2**). Patients with PBO recurrence exhibited significantly longer median tBCR compared with those with PLNI (37.5 versus 15 months, p=0.006). Seminal vesicle invasion (p=0.006) and advanced pT-stage (p=0.01) emerged as the most prominent pathological factor associated with nodal involvement. Univariable LR analysis identified SVI (OR, 5.31; 95% CI, 1.60-17.59; p=0.006) and advanced pT-stage (OR= 2.35; 95% CI, 1.24-4.46; p=0.009) as significant predictors of pelvic lymph node involvement. Variables demonstrating marginal associations include pN-stage (OR, 2.28; 95% CI, 0.99-5.28; p=0.053) and tBCR (OR, 0.43; 95% CI, 0.18-1.01; p=0.058). On multivariable analysis adjusting for all variables with p<0.05 (SVI, pT-stage) or p<0.10 (adding pN-stage, TBR), neither threshold yielded independently significant predictors.

**Table 2:** Clinicopathology stratified by anatomical site of recurrence.

| Variables | PBO | PLNI | p |
| --- | --- | --- | --- |
| N | 28 | 49 |  |
| Age (mean ± SD) | 69 ± 7.9 | 67.8 ± 7.2 | 0.30 |
| pISUP |  |  |  |
| 1 | 3 | 1 | 0.205 |
| 2 | 10 | 13 |  |
| 3 | 9 | 19 |  |
| 4 | 3 | 3 |  |
| 5 | 3 | 13 |  |
| pT-stage |  |  |  |
| T2 | 10 | 10 | 0.01 |
| T3a | 14 | 15 |  |
| T3b | 4 | 24 |  |
| pN-stage |  |  |  |
| Nx | 6 | 3 | 0.095 |
| N0 | 17 | 31 |  |
| N1 | 5 | 15 |  |
| EPE |  |  |  |
| Yes | 17 | 23 | 0.245 |
| No | 11 | 26 |  |
| PSM |  |  |  |
| Yes | 10 | 15 | 0.645 |
| No | 18 | 34 |  |
| SVI |  |  |  |
| Yes | 4 | 23 | 0.004 |
| No | 24 | 26 |  |
| tBCR (month)<br>(median; IQR) | 37.5 (21.3- 126.5) | 15 (6-49) | 0.006 |
| BCR-PSA (median; IQR) | 0.6 (0.38-3.0) | 0.93 (0.51-1.74) | 0.421 |

Abbreviations: EPE, extraprostatic extension; pT-stage, pathological T-stage; pN-stage, pathological nodal stage; SVI, seminal vesicle invasion; PSM, positive surgical margin; tBCR, time to biochemical recurrence; BCR-PSA, PSA level at biochemical recurrence prior PSMA-PET; p, p-value

### Salvage Radiotherapy Outcomes

Of 140 patients treated with sRT, 124 had complete time-to-event data and comprised the evaluable cohort for outcome analysis. Salvage ADT status was available for 116 of these patients, of whom 90 (77.6%) received ADT. ADT use did not differ significantly between PSMA-PET-positive and PSMA-PET-negative patients (51/64 [79.7%] vs. 38/52 [73.1%]; p=0.40). Median follow-up was 31 months (IQR; 21-41) with sBCR occurring in 33 of 124 patients (26.6%). Estimated bDFS at 12, 24, 36, and 60 months was 98.3%, 89.7%, 80.2%, and 54.1%, respectively **(Fig. 2)**.

**Fig 2:**
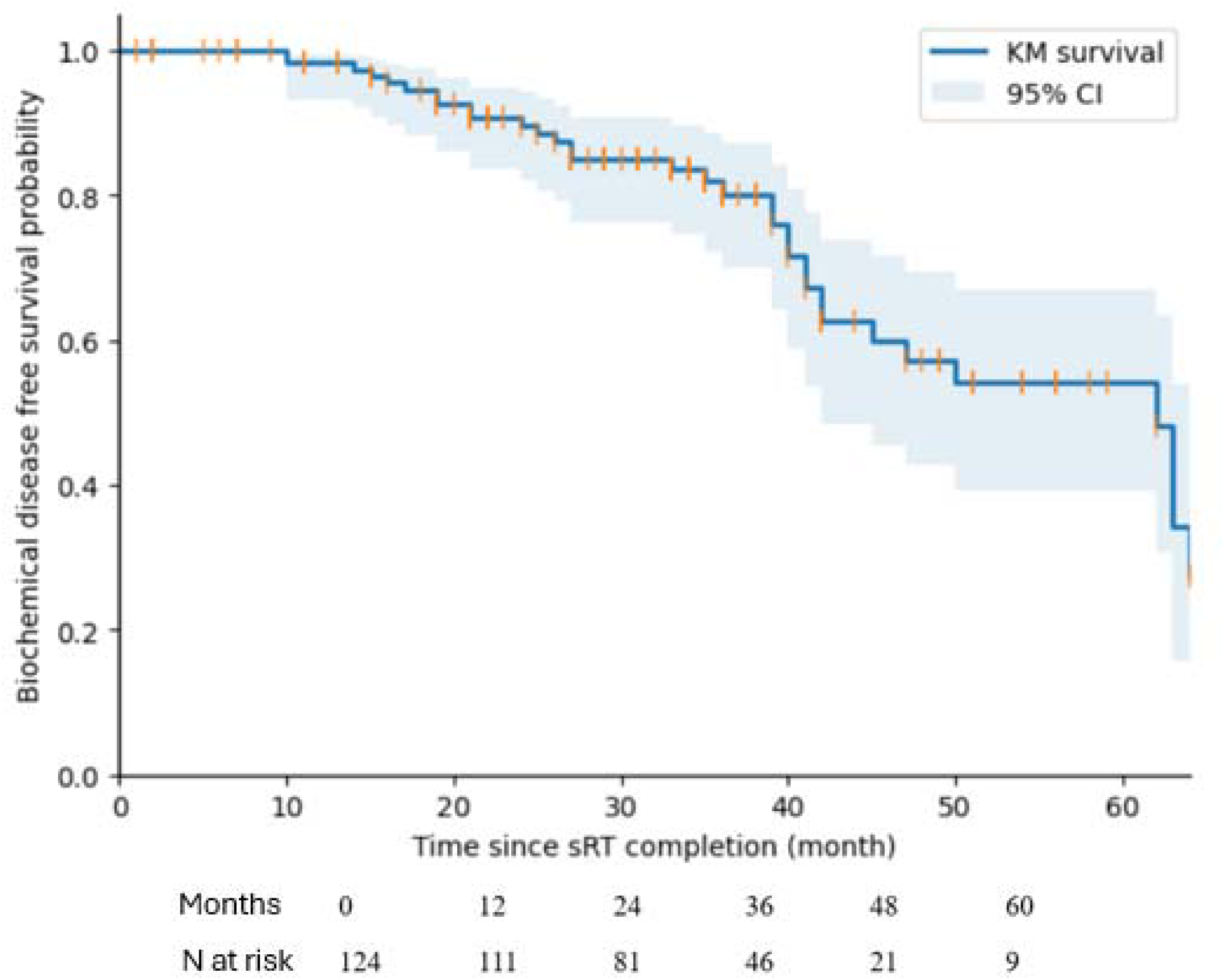
Kaplan-Meier estimates of biochemical disease-free survival following salvage radiotherapy. Shaded area represents 95% confidence interval. Tick marks indicate censored observations. Abbreviations: CI, confidence interval; KM, Kaplan-Meier; sRT, salvage radiotherapy; N, number of patients at risk.

Biochemical disease-free survival did not differ significantly between PSMA-PET-positive and PSMA-PET-negative patients across the entire follow-up period (sBCR rate 19/68 versus 14/56; log-rank p=0.775). Three-year bDFS estimates were 72.6% (CI 56.5-83.6) and 89.2% (75.73-95.38) for PSMA-PET-positive and PSMA-PET-negative patients, respectively, consistent with the overall non-significant log-rank result (**Fig. 3A**). Among PSMA-PET-positive patients with complete follow-up (n=68), bDFS did not differ significantly by recurrence pattern (sBCR rates: 5/23 (22%) for PBO versus 14/45 (31%) for PLNI; log-rank p=0.353). Three-year bDFS estimates were 90% (95% CI= 67-97.5) for PBO and 63.3% (95% CI= 42.14-78.52) for PLNI (**Fig. 3B**). Kaplan-Meier survival curves stratified by significant clinicopathology variables including SVI (log-rank p=0.005), EPE (log-rank p=0.008), and pT-stage (log-rank p=0.056) are shown in **Fig. 4**.

**Fig 3:**
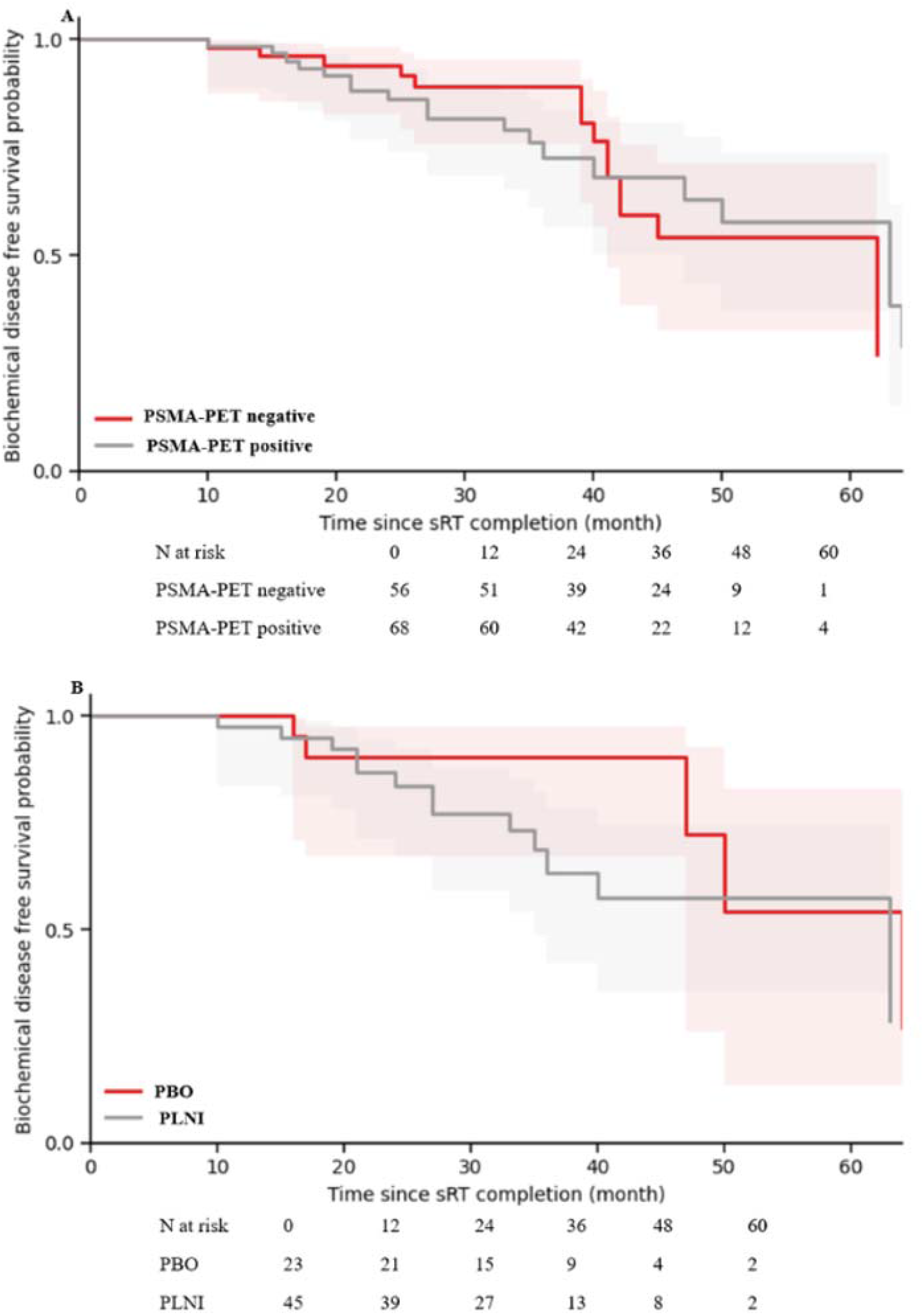
Biochemical disease-free survival stratified by (A) PSMA-PET findings and (B) site of anatomical recurrence patterns for PSMA-PET positive. Shaded areas represent 95% confidence intervals. Abbreviations: PBO, prostate bed-only recurrence; PLNI, pelvic lymph node involvement; sRT, salvage radiotherapy.

**Fig 4:**
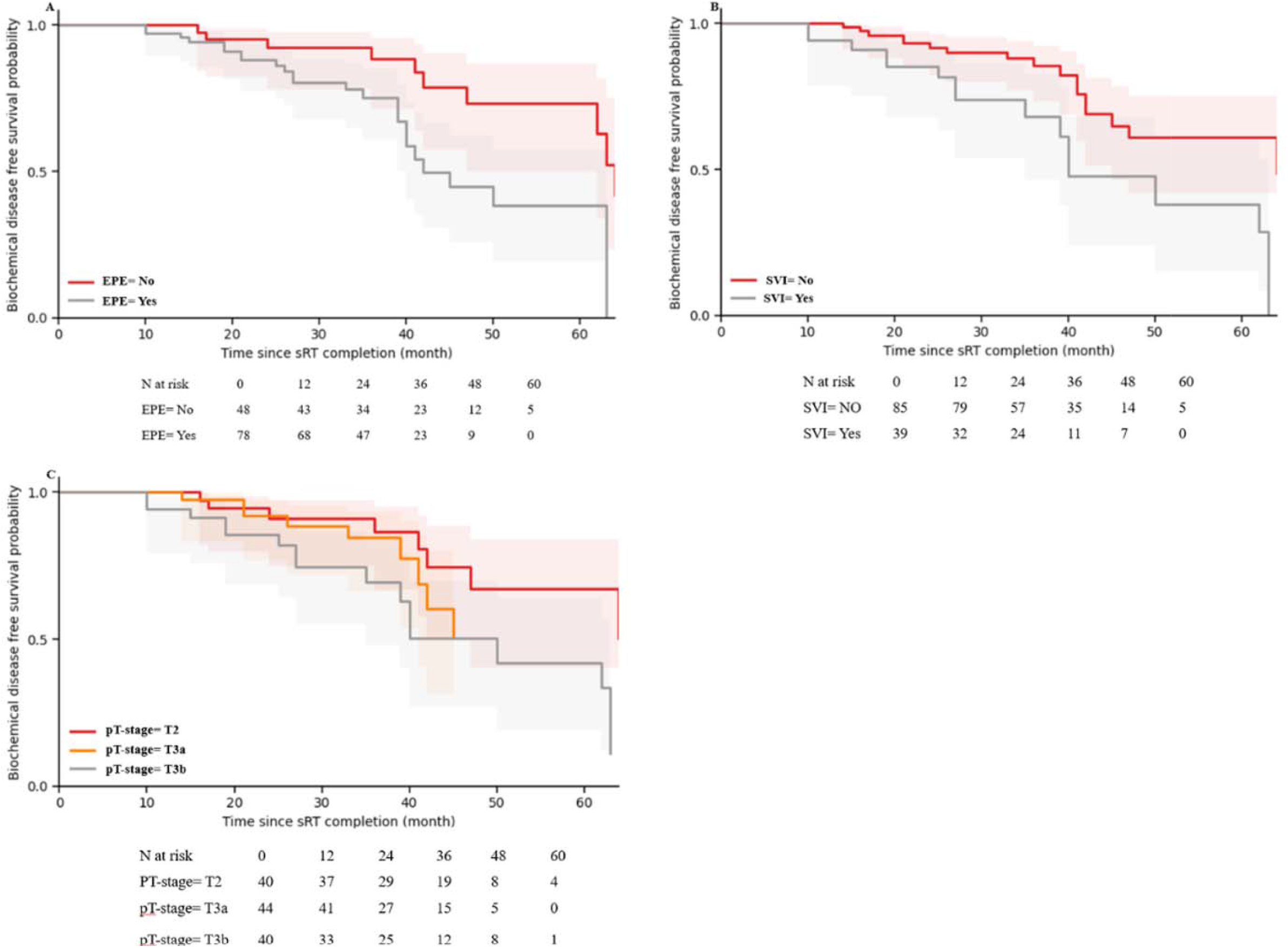
Biochemical disease-free survival stratified by adverse pathological features; (A). EPE (Yes versus No). (B). SVI (Yes versus No). (C). pT-stage (T2 versus T3a versus T3b). Shaded areas represent 95% confidence intervals. Abbreviation: sRT, salvage radiotherapy; EPE, extraprostatic extension; SVI, seminal vesicle invasion; pT-stage, pathology T-stage.

On univariable Cox proportional hazard analysis, SVI (HR=2.39, 95% CI=1.08-5.25; p=0.031), EPE (HR=2.82, 95% CI=1.26-6.32; p=0.012), pT-stage (HR=1.64, 95% CI=1.08–2.50; p=0.021), and pN-stage (HR=1.83, 95% CI=1.01-3.32; p=0.045) were associated with increased hazard of secondary biochemical failure. The multivariable Cox model incorporating all univariably significant covariates showed that none retained independent statistical significance, with wider confidence intervals.

## Discussion

This study provides a clinically integrated evaluation of how clinicopathologic characteristics influence PSMA-PET detectability, anatomical recurrence patterns, and biochemical outcomes following sRT in men with post-RP BCR. Three principal observations emerge. First, biochemical tumor burden reflected by BCR-PSA, together with patient age at time of recurrence, were the dominant independent determinants of PSMA-PET detection. Second, among the PSMA-PET patients with locoregional disease, adverse pathological features, particularly the presence of SVI and more advanced pT-stage, were associated with pelvic nodal recurrence patterns, but these associations were substantially diminished after multivariable adjustment. Third, neither clinicopathologic variables nor PSMA-defined locoregional recurrence patterns demonstrated an independent association with bDFS after sRT.

The locoregional detection rate observed in our cohort is consistent with the established performance of PSMA-PET in BCR, which demonstrates considerable heterogeneity across studies depending on PSA thresholds and patient risk ^7,8,16^. Meta-analyses of ^68^Ga-PSMA-11 PET/CT in the BCR setting have reported pooled detection rates ranging from 47% to 80%, with detection probability closely linked to PSA burden ^8,17^.

The emergence of BCR-PSA as the dominant independent predictor of PSMA-PET positivity aligns with the observed stepwise increase in detection rates and reflects the close relationship between circulating PSA levels and overall tumor burden ^18–20^. The observed ROC performance with an optimal BCR-PSA threshold of 0.58 ng/mL in our study further supports clinical suggestions for PSMA-PET imaging at PSA levels ≥0.5 ng/mL, where detection rates often exceed 60%, while remaining informative at lower PSA values in selected high-risk cases ^21^. Although our identified BCR-PSA threshold of 0.58 ng/mL corresponded to optimal discrimination for PSMA-PET positivity, this value should not be interpreted as a recommended threshold for delaying salvage treatment. Rather, it reflects the increasing probability of lesion detectability with rising tumor burden. Contemporary evidence supports consideration of sRT at substantially lower PSA levels, as earlier treatment is associated with better oncologic outcomes despite lower PSMA-PET detection rates ^2,22^. Consequently, the clinical utility of PSMA-PET lies not in postponing therapy until higher PSA levels are reached but in improving disease localization and treatment personalization across the spectrum of biochemical recurrence.

Age at BCR also remained independently associated with PSMA-PET positivity, although the magnitude of the effect was modest. While age-related biological differences, including cumulative mutational burden and androgen axis dynamics, may contribute, this association may also reflect referral patterns, time-to-imaging intervals, or residual confounding ^23^. By contrast, the adverse pathology features such as pT-stage and EPE lost significance after adjusting for PSA. In primary staging, studies have reported that adverse pathological features are associated with enhanced PSMA expression or detectability ^24,25^. At recurrence, however, our result suggests PSA as a real-time biomarker of viable tumor burden mediates much of the apparent pathological risk on PSMA-PET detectability, reinforcing PSA as the primary clinical determinant of molecular imaging yield.

Among PSMA-positive patients, pelvic nodal involvement predominated, with PLNI patients demonstrating significantly shorter time to BCR compared with those with PBO recurrence. Similar distributions have been reported in 18-F-DCFPyL PSMA-PET cohorts by Sigurdson et al., where only a minority (18%) demonstrated isolated prostate bed recurrence, challenging earlier assumptions from pre-PSMA eras that post-RP recurrence was predominantly local, supporting the concept that occult regional dissemination is common even within locoregional boundaries ^26,27^. Seminal vesicle invasion and advanced pT-stage were both associated with nodal recurrence, supporting their role as markers of advanced local extension and lymphotropic spread, features known to correlate with adverse genomic signatures and metastatic potential ^28,29^. However, the absence of independent predictors on multivariable models likely suggests that once molecular imaging identifies disease, traditional pathological features provide limited additional discrimination capacity for anatomical distribution.

Salvage radiotherapy remains the standard of care for a locoregional treatment approach for men with BCR after RP, particularly when recurrence is not overtly metastatic and treatment is delivered early ^4^. Consistent with our study, sRT series, including randomized trials GETUG-AFU 16 and RTOG 0534 SPPORT, demonstrates a 5-year biochemical disease control rate in the order of 52-80% ^30–32^. Although direct comparisons remain challenging given heterogeneity in patient selection, pre-salvage PSA levels, treatment delivery approach, ADT utilization, and outcome definitions.

Secondary biochemical failure occurred at a comparable rate between PSMA-positive and PSMA-negative patients, with no significant differences in bDFS according to PSMA status or recurrence patterns among PSMA-positive men. These findings do not imply lack of prognostic relevance for PSMA-PET; rather, they likely reflect the competing effects of PSMA-PET-guided treatment intensification. PSMA-PET-visible disease may signal greater macroscopic tumor burden or aggressive biology but enables targeted interventions, including expanded radiation fields, elective nodal irradiation, focal dose escalation to PET-avid lesions that partially offset adverse biological implications, as demonstrated in the study by Schmidt-Hegemann et al. ^6^. Differential androgen suppression could similarly narrow this comparison, as more intensive ADT use may delay PSA-defined biochemical failure partly through PSA suppression, rather than solely through improved durable disease control. However, ADT utilization did not differ significantly between PSMA-PET-positive and PSMA-PET-negative patients in our cohort, indicating that a gross imbalance in ADT use is unlikely to be the principal driver ^33^. By contrast, Emmett et al. demonstrated that PSMA-PET is independently predictive of treatment response to sRT, stratifying men into high-response (negative or fossa-confined PSMA) versus low-response groups (nodes or distant-disease PSMA) ^34^. Their divergence from our study is likely explained by their broader inclusion of metastatic disease, whereas our analysis was confined to the locoregional recurrence pattern among salvage-treated patients.

PSMA-negative BCR, while undetected by imaging, likely represents microscopic disease that yielded to standard prostate-bed irradiation, consistent with long-standing sRT data demonstrating durable control in appropriately selected patients in pre-molecular era ^5,35^. Conversely, PSMA-positive locoregional disease, despite a higher macroscopic burden, may benefit from intensification strategies that offset its biology ^6^. This framework is further supported by patterns-of-failure analyses indicating that post-sRT recurrence frequently arises at distant sites rather than within treated fields, suggesting that both locoregional disease, whether image-detected or microscopic, often serves as a surrogate for systemic burden ^36^. This highlights the need for composite risk models incorporating both imaging and pathological features as a prognostic stratifier for sRT outcomes.

In the pre-PSMA era, adverse pathological features such as the presence of SVI and positive surgical margin status have been identified as predictors of PSA progression after sRT ^37^. In our PSMA-PET-guided sRT cohort, these features, including SVI, EPE, advanced pT-stage, and pN-stage, retained univariable associations with bDFS; however, none demonstrated independent significance on multivariable analysis. This loss of independent prognostic value does not negate their biological importance but highlights challenges in isolating independent contributions in modern, imaging-informed salvage settings. The high frequency of pelvic nodal involvement among PSMA-positive patients supports routine consideration of elective pelvic nodal irradiation, particularly in early recurrence or adverse pathology (SVI, pT3b, pN1 disease). Close surveillance remains warranted for men with EPE, SVI, or advanced pathology stage post-prostatectomy, as these indicate greater tumor volume and occult micrometastatic risk, though prospective data are needed to validate treatment escalation ^38,39^.

This study benefits from several strengths, including comprehensive clinicopathological annotation, standardized outcome definitions, and inclusion of both PSMA-positive and PSMA-negative patients, enabling unbiased assessment of detection determinants and treatment efficacy across the full spectrum of biochemical recurrence. However, the study is limited by its retrospective single-center design and moderate sample size, limiting statistical power for multivariable survival modeling, particularly given the modest number of secondary biochemical failures and high collinearity among adverse pathologic features. Treatment-related variables, including ADT duration, extent of pelvic nodal irradiation field, and focal boost strategies, could not be fully incorporated into outcome models, introducing potential residual confounding. Time-to-event analyses were restricted to patients with adequate longitudinal follow-up, which may have introduced loss to follow-up bias.

## Conclusion

PSA level at biochemical recurrence and age independently predicted PSMA-PET detection, with pathological features contributing minimally. Among imaging-positive patients, pelvic nodal involvement predominated, associated with earlier recurrence and SVI/pT-stage on univariable analysis. sRT outcome in patients with locoregional recurrence remained independent of PSMA-PET status or recurrence pattern, yielding comparable bDFS across strata. This highlights the biological heterogeneity of recurrent prostate cancer and supports prospective validation to optimize risk-adapted salvage intensification.

## Author Contributions

Ramotallah Jubril contributed to the study design, collected and analyzed data, and drafted the first version of the manuscript. Oluwaseyi Oderinde designed the study and revised the manuscript. Arpan V. Prabhu, Omar Ishaq, Jesus Ocana, Mark Tann, Christopher Njeh, Clinton Bahler, and Jordan Holmes participated in the analysis and interpretation of data and commented on the previous version of the manuscript. All authors read and approved the final manuscript.

## Data Availability

All data generated during and/or analyzed in this study are available from the corresponding author upon reasonable request.

## Ethics Statement

This study was approved by the Institutional Review Board of Indiana University (IRB No. 11330), and all study protocols were performed in accordance with the principles of the Declaration of Helsinki. Due to the retrospective nature of the data collection, formal written patient consent was waived.

